# Association between human blood metabolome and the risk of coronary heart disease: Mendelian randomization study

**DOI:** 10.1101/2024.01.31.24302109

**Authors:** Jia Zhu, Xiaojun Xia, Haodong Jiang, Congying Wang, Yunpeng Jin

## Abstract

In this study, we employed Mendelian Randomization (MR) to elucidate the causal relationships between specific blood metabolites and Coronary Heart Disease (CHD). By analyzing data from Genome-Wide Association Studies (GWAS) and the FinnGen database, we conducted a two-sample MR analysis focusing on 40 metabolites and 6 metabolite ratios linked to CHD risk. Our findings highlight a group of metabolites significantly influencing CHD risk, either augmenting or mitigating it. Rigorous sensitivity checks, including MR-Egger and MR-PRESSO, negated the influence of horizontal pleiotropy and reinforced the robustness of our results. Furthermore, reverse MR analysis unveiled a bidirectional influence between certain metabolites and CHD, suggesting a complex mutual interaction. This study not only unravels intricate connections between metabolites and CHD, but also paves the way for potential biomarkers instrumental in CHD prevention and therapy. However, it acknowledges certain limitations, such as the modest sample size and a primary focus on European genetic data, underscoring the need for further investigations in more diverse population cohorts.

## Introduction

Coronary Heart Disease (CHD) is a complex chronic inflammatory disorder, characterized predominantly by the narrowing and remodeling of coronary arteries. Despite comprehensive research into the pathogenesis of CHD^1, 2^, leading to numerous therapeutic approaches for coronary artery disease^3^, recent epidemiological studies indicate that CHD remains a leading cause of mortality^4^. Therefore, understanding new mechanisms in the development and progression of CHD is crucial for its prevention and treatment^5, 6^.

Metabolites in the blood, reflective of physiological state changes, are increasingly recognized for their role in the early detection and prognosis of CHD^7-9^. However, due to confounding factors and reverse causation, the causal relationship between human metabolites and CHD remains to be fully elucidated.

Employing Mendelian Randomization (MR), a sophisticated epidemiological approach, which offers a more convenient means of eliminating confounders and verifying causal relationships compared to Randomized Controlled Trials (RCTs)^10-12^, this study utilizes two-sample MR analysis. Utilizing the latest Genome-Wide Association Studies (GWAS) and the FinnGen database, this analysis investigates the causal relationship between exposures (metabolites) and outcomes (CHD), providing potential targets for subsequent CHD interventions.

## Materials and methods

### Study design

In this study, a two-sample Mendelian Randomization (MR) framework is employed to rigorously evaluate the causal links between specific metabolites and Coronary Heart Disease (CHD), as delineated in Figure 1. Within the ambit of MR analysis, the instrumental variables (IVs), constituted by Single Nucleotide Polymorphisms (SNPs)13, are mandated to fulfill three stringent criteria: 1. The selected SNPs, serving as IVs, must exhibit a robust association with the exposure, evidenced by an F-statistic >10. 2. These SNPs should demonstrate no association with the outcome variable. 3. The SNPs must remain uncorrelated with any potential confounding variables, thereby ensuring the integrity and validity of the causal inferences drawn.14

**Figure 1.**
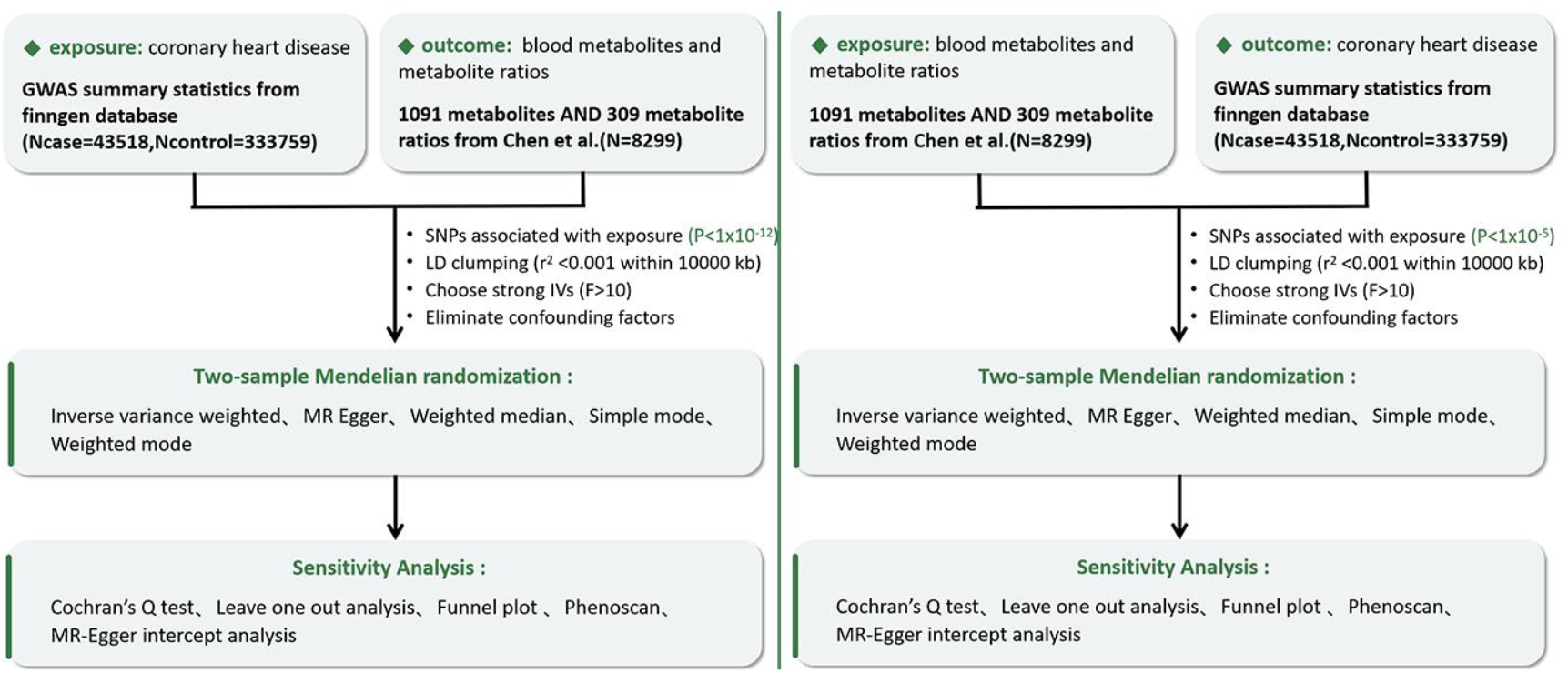
Overview of this MR study

### Genome-wide association study(GWAS)data sources for blood metabolites and CHD

Genetic variations related to metabolite levels were derived from an extensive Genome-Wide Association Study (GWAS) within the Canadian Longitudinal Study on Aging (CLSA) cohort, analyzing 1091 types of metabolites and 309 metabolite ratios^15^. The GWAS data for CHD originated from the Finnish database (N_case_=43518, N_control_=333759).

### Selection of instrumental variables

In this study, we identified a set of Single Nucleotide Polymorphisms (SNPs) associated with blood metabolites, each surpassing the genome-wide significance threshold (P-value < 1×10^-5)^16, 17^, thereby qualifying them as instrumental variables (IVs)^18^. This identification was achieved through a comprehensive and sequential filtration process

Firstly, employing PLINK software (version v1.90), we executed a clumping procedure to refine the SNPs selection. This process adhered to a strict linkage disequilibrium (LD) r^2 threshold of less than 0.001, within a 10,000 kb window,^19^ ensuring the minimization of confounding genetic linkage. Secondly, to enhance the robustness of our IVs, we systematically excluded those with weaker statistical relevance, particularly focusing on those with an F-statistic < 10.^20^ This stringent selection criterion was pivotal in reinforcing the reliability of our instrumental variables.

Finally, to address and mitigate potential confounders specifically pertaining to Coronary Heart Disease (CHD), an exhaustive search was conducted using the Phenoscanner database (http://www.phenoscanner.medschl.cam.ac.uk/).^21^ Any IVs found to be associated with these confounders were rigorously excluded from our analysis, thereby refining our assessment and ensuring the integrity of our causal inferences

### Statistical analysis

The association between metabolites and Coronary Heart Disease (CHD) underwent a rigorous evaluation through the application of the ‘Mendelian-Randomization’ package (Version 0.4.3)^22^. Our analysis leveraged advanced statistical methods including inverse variance weighting (IVW), weighted median^23^, and mode-based estimation, facilitating a multifaceted assessment. To enhance the reliability of our results, we meticulously computed Cochran’s Q statistic^24^ accompanied by corresponding p-values, offering a comprehensive quantification of heterogeneity across the employed instrumental variables (IVs).

Concurrently, we utilized the MR-EAGGER methodology^25^, renowned for its efficacy in identifying pleiotropy via significant intercepts, to effectively address horizontal pleiotropy within our dataset. We also implemented a leave-one-out analysis to further validate the robustness of our findings. Complementary to these quantitative methods, our study integrated detailed scatter plots and funnel charts, meticulously constructed to provide a visual interpretation of our data, thereby underscoring the validity of our analytical approach^26^

### Reverse MR analysis

In our analysis, we adopted a more stringent threshold (p < 5×10^-12) for selecting Single Nucleotide Polymorphisms (SNPs) associated with Coronary Heart Disease (CHD) from the FinnGen database. Subsequently, the filtered Instrumental Variables (IVs) underwent a similar selection process as initially employed for the metabolite-associated SNPs.

## Results

### Levels of blood metabolites associate with CHD risk

To investigate the influence of blood metabolites and changes in their ratios on coronary heart disease (CHD), we conducted a two-sample Mendelian Randomization (MR) analysis using the Inverse Variance Weighted (IVW) method as our primary approach. Our study identified 46 metabolites and 7 metabolite ratios impacting CHD risk. After adjusting for the False Discovery Rate (FDR), we excluded results with PFDR > 0.05. This exclusion applied to the Adenosine 5′-monophosphate (AMP) to palmitate (16:0) ratio (P_FDR_=0.0568), along with levels of 3beta-hydroxy-5-cholestenoate (P_FDR_=0.0513), Cystathionine (P_FDR_=0.0523), Decanoylcarnitine (C10) (P_FDR_=0.5), Methylsuccinoylcarnitine (P_FDR_=0.527), Ornithine (P_FDR_=0.527), and 2-hydroxyhippurate (salicylurate) (P_FDR_=0.526). Ultimately, our analysis confirmed that 40 metabolites and 6 metabolite ratios are significant influencers of CHD risk. Of these, 24 metabolites and 3 metabolite ratios appear to increase CHD risk, while 16 metabolites and 3 ratios seem to offer a protective effect. (Figure 2 and Supplementary Figure 1) The outcomes presented herein have undergone comprehensive global tests through both MR-Egger and MR-PRESSO methodologies, effectively ruling out the presence of horizontal pleiotropy. Additionally, scatter plots and funnel plots have been constructed to demonstrate the stability and robustness of the results.

**Figure 2.**
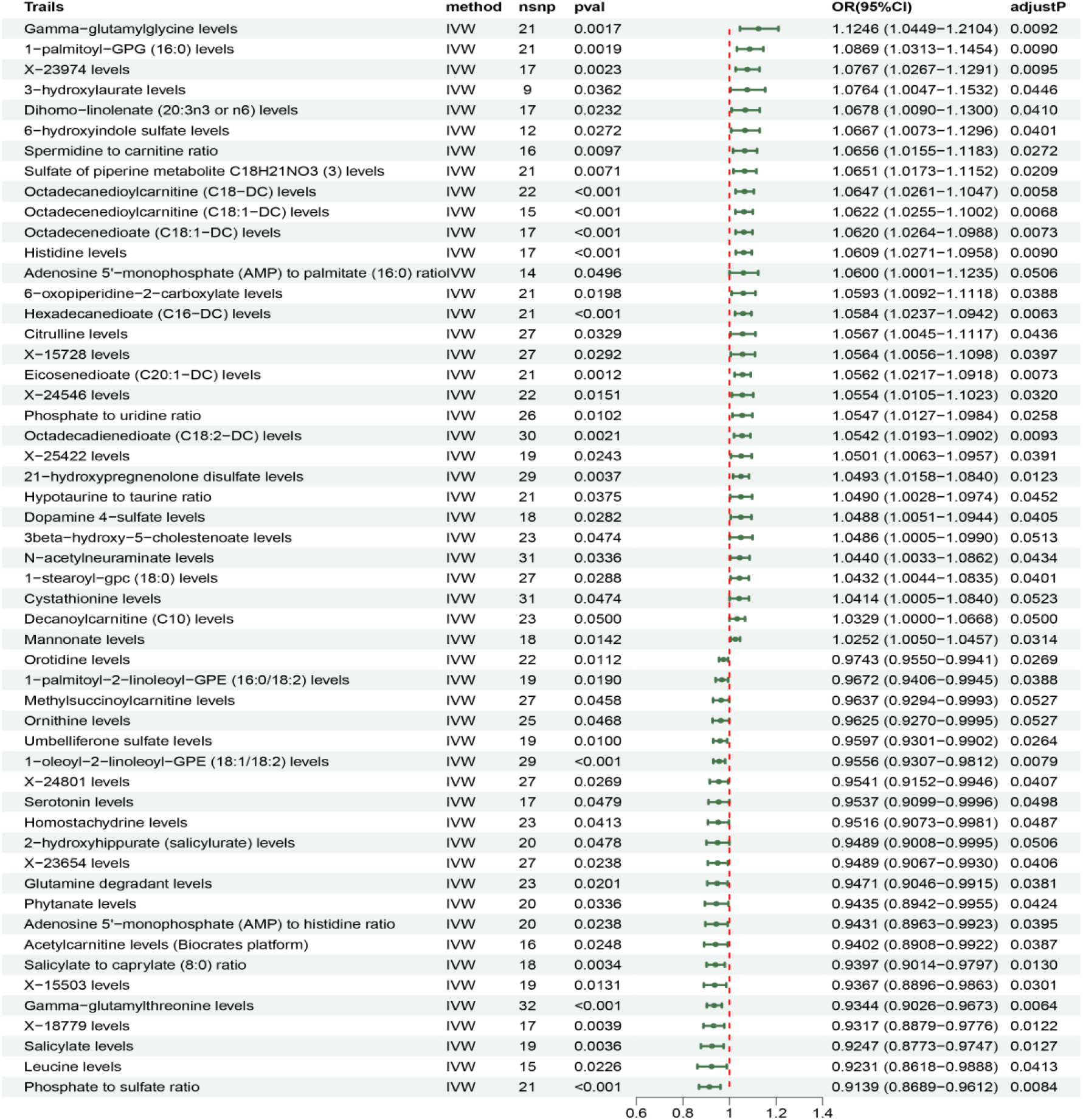
Forest plots showed the causal associations metabolites and metabolites ratio and CHD

### Reverse MR analysis assessing the effect of CHD on LEVELS Of blood metabolities

To investigate the variations in metabolites and their ratios associated with the onset of coronary heart disease (CHD), we again employed a two-sample Mendelian Randomization (MR) analysis. After comprehensive adjustments for the False Discovery Rate (FDR), we found that CHD onset is characterized by increased levels of four specific metabolites: Dihomo-linolenate (20:3n3 or n6) (P_FDR_=0.0452), 2-hydroxyhippurate (salicylurate) (P_FDR_=0.0035), Salicylate (P_FDR_=0.0018), and X-15503 (P_FDR_=0.0186).(Figure 3 and Supplementary Figure 2) The same sensitivity analyses have corroborated the stability of the observed outcomes. The scatter plots and funnel plots further substantiate the reliability of these results.

**Figure 3.**
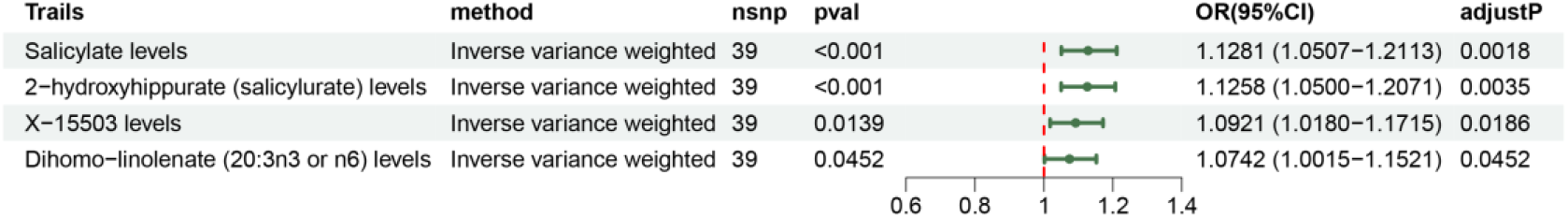
Forest plots showed the causal associations CHD and metabolites and metabolites ratio

## Discussion

Through an exhaustive analysis of extensive genetic data, this research uncovers bidirectional causal relationships between alterations in blood metabolites and their ratios, and the risk of coronary heart disease (CHD), as elucidated via Mendelian Randomization (MR) analysis. The study discerns 40 metabolites and 6 metabolite ratios significantly correlated with CHD risk (P<0.05). A reverse causation analysis reveals an elevation in four blood metabolites subsequent to CHD onset. Augmented levels of 2-hydroxyhippurate (salicylurate), Salicylate, and X-15503 are inversely associated with CHD risk, implying a potential protective role under normative conditions. However, the emergence of CHD seems to instigate a compensatory increase in these metabolites, elucidating a multifaceted interaction. This observation is congruent with prior research indicating stress responses triggered by CHD, leading to elevated metabolite levels. In contrast, an escalation in Dihomo-linolenate (20:3n3 or n6) levels is connected to an increased risk of CHD, with further elevation post-CHD onset, suggesting a bidirectional relationship and hinting at an intricate network of interactions or shared etiological factors.

Employing the latest GWAS summary data, this study eclipses previous research in terms of scale of exposure data (8299 compared to 7828) and a wider range of metabolites^27^, ensuring a more thorough and rigorous analysis. While the selection criteria were exacting, more lenient conditions were adopted for identifying factors influencing CHD (genome-wide significance level of p<1× 10^-5, r2<0.001 within 10000kb). For the reverse causation analysis, owing to the broader conceptualization of CHD relative to CAD and the availability of additional instrumental variables, more stringent criteria were applied (genome-wide significance level of 1×10^−12^, r2<0.001 within 10000kb). The presence of multiple instrumental variables (IVs) for the metabolites necessitated a range of sensitivity analyses, which did not reveal any significant pleiotropy or heterogeneity.

Nonetheless, the study is subject to certain limitations. The relatively modest sample size for the selected metabolites might introduce bias. Furthermore, the exclusive reliance on genetic data from European populations curtails the universality of the findings. Future research is warranted in more diverse populations to enhance the generalizability of these relationships. The insights gleaned from this study pave the way for novel biomarkers and potential targets in CHD prevention and treatment strategies, offering new perspectives in the realm of cardiovascular disease research.

## Conclusion

Our research reveals that alterations in 40 metabolites and 6 metabolite ratios can influence the occurrence of coronary heart disease (CHD). These findings suggest a complex network of interactions between them and CHD, offering novel insights for understanding the mechanisms of CHD progression, and paving the way for improved prediction, prevention, and treatment strategies.

## Acknowledgements

This study was possible thanks to publicly available GWAS summary statistics, including those from the GWAS catalog and IEU OpenGWAS project.

## Author contributions

J.Z. performed the research, analyzed the data, and wrote the manuscript. X.-J.X. and H.-D.J. draw the figures. C.-Y.W. wrote a part of the manuscript. Y.-P.J. revised, and approved the final manuscript and is the corresponding author. All authors read and approved the final version of the manuscript.

## Data availability

The summary statistics of SNP-metabolite associations from the CLSA study were available from the NHGRI-EBI GWAS Catalog (https://www.ebi.ac.uk/gwas/) with accession number GCST90199621-90201020. The summary statistics of CHD GWAS were available from the Finngen(https://www.finngen.fi/en)

## References

1. Stone PH, Libby P, Boden WE. Fundamental Pathobiology of Coronary Atherosclerosis and Clinical Implications for Chronic Ischemic Heart Disease Management-The Plaque Hypothesis: A Narrative Review. JAMA cardiology. 2023;8: 192–201.

2. Libby P, Buring JE, Badimon Let al. Atherosclerosis. Nature reviews Disease primers. 2019;5: 56.

3. Dibben GO, Faulkner J, Oldridge Net al. Exercise-based cardiac rehabilitation for coronary heart disease: a meta-analysis. European heart journal. 2023;44: 452–469.

4. Dalen JE, Alpert JS, Goldberg RJ, Weinstein RS. The epidemic of the 20(th) century: coronary heart disease. The American journal of medicine. 2014;127: 807–812.

5. Lim SY, Selvaraji S, Lau H, Li SFY. Application of omics beyond the central dogma in coronary heart disease research: A bibliometric study and literature review. Computers in biology and medicine. 2022;140: 105069.

6. Pouralijan Amiri M, Khoshkam M, Salek RM, Madadi R, Faghanzadeh Ganji G, Ramazani A. Metabolomics in early detection and prognosis of acute coronary syndrome. Clinica chimica acta; international journal of clinical chemistry. 2019;495: 43–53.

7. Dong Y, Chen H, Gao J, Liu Y, Li J, Wang J. Molecular machinery and interplay of apoptosis and autophagy in coronary heart disease. Journal of molecular and cellular cardiology. 2019;136: 27–41.

8. Talmor-Barkan Y, Bar N, Shaul AAet al. Metabolomic and microbiome profiling reveals personalized risk factors for coronary artery disease. Nature medicine. 2022;28: 295–302.

9. Hu J, Yao J, Deng Set al. Differences in Metabolomic Profiles Between Black and White Women and Risk of Coronary Heart Disease: an Observational Study of Women From Four US Cohorts. Circulation research. 2022;131: 601–615.

10. Davey Smith G, Hemani G. Mendelian randomization: genetic anchors for causal inference in epidemiological studies. Human molecular genetics. 2014;23: R89–98.

11. Larsson SC, Butterworth AS, Burgess S. Mendelian randomization for cardiovascular diseases: principles and applications. European heart journal. 2023;44: 4913–4924.

12. Lawlor DA, Harbord RM, Sterne JA, Timpson N, Davey Smith G. Mendelian randomization: using genes as instruments for making causal inferences in epidemiology. Statistics in medicine. 2008;27: 1133–1163.

13. Sheehan NA, Didelez V. Epidemiology, genetic epidemiology and Mendelian randomisation: more need than ever to attend to detail. Human genetics. 2020;139: 121–136.

14. Luo J, le Cessie S, van Heemst D, Noordam R. Diet-Derived Circulating Antioxidants and Risk of Coronary Heart Disease: A Mendelian Randomization Study. Journal of the American College of Cardiology. 2021;77: 45–54.

15. Chen Y, Lu T, Pettersson-Kymmer Uet al. Genomic atlas of the plasma metabolome prioritizes metabolites implicated in human diseases. Nature genetics. 2023;55: 44–53.

16. Auton A, Brooks LD, Durbin RMet al. A global reference for human genetic variation. Nature. 2015;526: 68–74.

17. Dai N, Deng Y, Wang B. Association between human blood metabolome and the risk of hypertension. BMC genomic data. 2023;24: 79.

18. Burgess S, Small DS, Thompson SG. A review of instrumental variable estimators for Mendelian randomization. Statistical methods in medical research. 2017;26: 2333–2355.

19. Wang C, Zhu D, Zhang Det al. Causal role of immune cells in schizophrenia: Mendelian randomization (MR) study. BMC psychiatry. 2023;23: 590.

20. Pierce BL, Ahsan H, Vanderweele TJ. Power and instrument strength requirements for Mendelian randomization studies using multiple genetic variants. International journal of epidemiology. 2011;40: 740–752.

21. Kamat MA, Blackshaw JA, Young Ret al. PhenoScanner V2: an expanded tool for searching human genotype-phenotype associations. Bioinformatics (Oxford, England). 2019;35: 4851–4853.

22. Yavorska OO, Yavorska OO, Burgess S, Burgess SJIJoE. MendelianRandomization: an R package for performing Mendelian randomization analyses using summarized data. 2017;46: 1734–1739.

23. Bowden J, Davey Smith G, Haycock PC, Burgess S. Consistent Estimation in Mendelian Randomization with Some Invalid Instruments Using a Weighted Median Estimator. Genetic epidemiology. 2016;40: 304–314.

24. Egger M, Smith GD, Phillips AN. Meta-analysis: principles and procedures. BMJ (Clinical research ed). 1997;315: 1533–1537.

25. Burgess S, Bowden J, Fall T, Ingelsson E, Thompson SG. Sensitivity Analyses for Robust Causal Inference from Mendelian Randomization Analyses with Multiple Genetic Variants. Epidemiology (Cambridge, Mass). 2017;28: 30–42.

26. Verbanck M, Chen CY, Neale B, Do R. Detection of widespread horizontal pleiotropy in causal relationships inferred from Mendelian randomization between complex traits and diseases. Nature genetics. 2018;50: 693–698.

27. Qiao J, Zhang M, Wang T, Huang S, Zeng P. Evaluating Causal Relationship Between Metabolites and Six Cardiovascular Diseases Based on GWAS Summary Statistics. Frontiers in genetics. 2021;12: 746677.

